# Personal protective equipment does not sufficiently protect against virus aerosol unless combined with advanced air purification or ventilation techniques

**DOI:** 10.1101/2021.09.02.21263008

**Authors:** Shane A Landry, Dinesh Subedi, Jeremy J Barr, Martin I MacDonald, Samantha Dix, Donna M Kutey, Darren Mansfield, Garun S Hamilton, Bradley A. Edwards, Simon A Joosten

## Abstract

**Background:** Healthcare workers (HCWs) are at risk from nosocomial transmission of SARS-CoV-2 from virus laden aerosols. This study aimed to: 1) quantify the degree of protection from virus aerosol provided by different types of mask (surgical, N95, fit-tested N95) and personal protective equipment (PPE); 2) determine if the use of a portable HEPA filter can enhance the effectiveness of PPE; 3) determine the effectiveness of a decontamination shower to remove virus aerosol contamination of a HCW.

**Methods:** Virus aerosol exposure experiments were conducted using bacteriophage PhiX174 (10^8^copies/mL). A HCW wearing PPE (mask, gloves, gown, faceshield) was exposed to nebulised viruses for 40mins in a sealed clinical room. After exiting, the HCW doffed PPE. Virus exposure was quantified via skin swabs applied to the face and nostrils, forearms, neck, and forehead. Experiments were performed with and without the presence of a portable HEPA filter (set to 470m3/hr).

**Findings:** Swabs quantified significant virus exposure under the surgical and N95 mask. Only the fit-tested N95 resulted in lower virus counts compared to no mask control (p=0.027). Nasal swabs demonstrated very high virus exposure, which was not mitigated by the surgical or N95 masks, although there was a trend for the fit-tested N95 mask to reduce virus counts (p=0.058). The addition of HEPA filtration substantially reduced virus counts from all swab sites, and to near zero levels when combined with a fit-tested N95 mask, gloves, gown and faces shield. Virus counts were substantially reduced to near zero levels following a shower.

**Interpretation:** These data demonstrate that quantitatively fit tested N95 masks combined with a HEPA filter can offer protection against high virus aerosol loads at close range and for prolonged periods of time. Skin contamination from virus aerosol can be effectively by removed by showering.

**Funding:** Epworth Hospital Capacity Building Research Grant ID: EH2020-654

## INTRODUCTION

The World Health Organization (WHO) and Centres for Disease Control and Prevention (CDC) updated their advice regarding the critical role of airborne transmission of SARS-CoV-2^1,2^ in April and May of 2021, respectively. Both organisations report that virus laden aerosols can remain suspended in the air for prolonged periods of time and travel large distances whilst remaining infectious. A critical modifying risk factor for aerosol transmission of SARS-CoV-2 is poorly ventilated spaces, which promote the accumulation of infectious aerosol.

Epidemiological evidence from previous sudden acute respiratory syndrome (SARS) outbreaks of SARS-CoV infections pointed to the importance of aerosol transmission.^3,4^ Nosocomial infection risk was demonstrated to be greatest in the setting of so-called aerosol generating procedures^5^, which resulted in widespread guidance for enhanced respiratory protection (personal and environmental) for healthcare workers (HCWs) caring for COVID-19 patients undergoing such procedures. However, recent work has shown that HCWs caring for patients not receiving aerosol generating procedures were contracting COVID-19 despite the use of surgical masks and personal protective equipment (PPE)^6–8^. Subsequent studies demonstrated that aerosols laden with infectious SARS-CoV-2 virus particles are present in the rooms of COVID-19 patients in the absence of aerosol generating procedures^9^ because aerosols are self-generated by COVID-19 patients when they cough, talk and breathe.

The above findings led the CDC to change their guidelines to now recommend that HCWs caring for COVID-19 patients wear an N95 or equivalent respirator and that indoor air quality is optimised including the deployment of portable high efficiency particulate absorbing (HEPA) filters when enhancement of permanent air-handling systems is not feasible.^10^ However, the rank importance of these measures is not clear and the magnitude of effect on infection risk reduction is not known. There is evidence that each measure in isolation (enhanced personal protection and enhanced environmental protection) may be inadequate. For example, pre-COVID studies demonstrated that respiratory infections still occur in the context of N95^11^ and fit tested N95 respirators^12^. Furthermore, a Cochrane database review showed uncertainty for any benefit of N95 over surgical mask in protecting against respiratory illness or lab confirmed influenza.^13^ With regard to environmental protection, COVID-19 can still be transmitted in outdoor settings^14^ and airborne SARS-CoV-2 can still be detected in negative pressure isolation rooms (12 air exchanges/hour) of patients with COVID-19.^15^ No study has systematically examined the interaction between personal and environmental protection in protecting against contamination with live virus aerosol.

We aimed to examine three mechanistic questions in relation to the effectiveness of personal PPE and air filtration to protect HCWs against virus aerosol contamination. Firstly, we aimed to quantify the degree of personal contamination with virus aerosol when wearing different types of mask (surgical, poor fitting N95, quantitatively fit tested N95) in combination with faceshield, gown and gloves. Secondly, we aimed to determine if the use of a portable HEPA filter enhanced the effectiveness of PPE to protect the wearer against virus aerosol contamination. Thirdly, we aimed to determine the effectiveness of a decontamination shower protocol to effectively remove virus aerosol contamination from the skin and upper airway of a HCW.

## METHODS

### Bacteriophage PhiX174 propagation and titration

Bacteriophage PhiX174 was used as a model virus in all experiments. PhiX174 was propagated using bacterial host *Escherichia coli* C (ATCC 13706) in lysogeny broth. The bacteriophage was purified according to the Phage-on-Tap protocol^16^. A titre of 1-5×10^9^ plaque-forming units (PFU)/mL was obtained and diluted as required in 1X phosphate-buffered saline (PBS; Omnipur, Gibbstown, NJ, USA). The bacteriophage titre was determined using the standard soft agar overlay method. A 10ml bacteriophage lysate (10^8^copies/mL) was aerosolised in all experiments.

### Simulated virus aerosol exposure

A nebuliser (PARI Respiratory Equipment, VA, USA), positioned at the head of a clinical bed, was used to aerosolise the bacteriophage lysate within a simulated clinical room (dimensions: 4.0 × 3.25 × 2.7 m, volume = 35.1 m^3^, see Figure 1). The Pari-PEP nebulizer produces a distribution of aerosol particle size of 3.42±0.15 µm^17^. We recorded particle mass concentration with a PurpleAir PA-II-SD (PurpleAir Inc. Utah, USA) sensor for reference (see supplement Figure S1)

**Figure 1.**
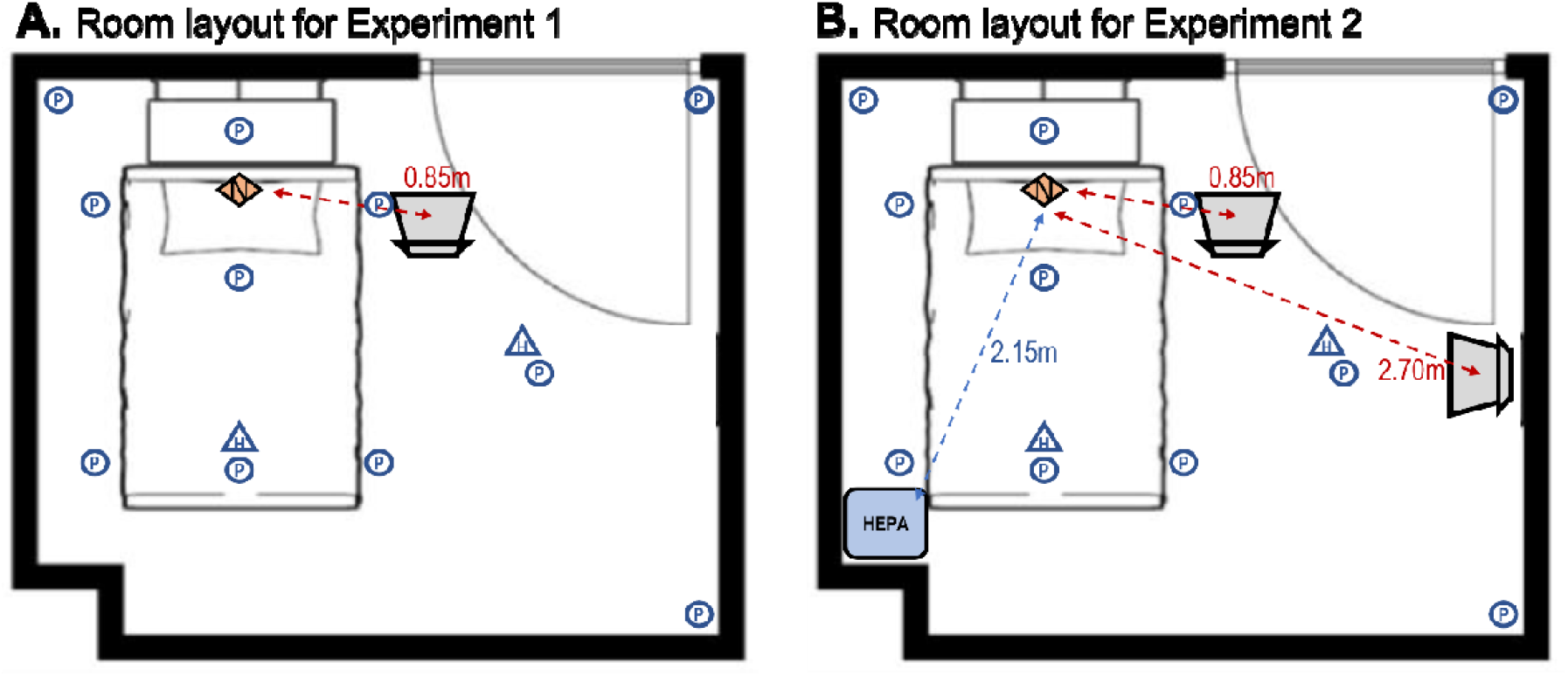
Experimental layout for experiments 1 and 2. All experiments were performed in a clinical room with dimensions 4.0 × 3.25 × 2.7 m (volume = 35.1 m^3^) containing a bed and one chair. Eleven settle plates (blue circles) were positioned identically between both experiments. Two hanging plates (blue triangles) were hung at head height perpendicular to the floor. The nebuliser (orange diamond) was positioned at the head of the bed, with the exit point facing vertically. **A)** In experiment 1 the HCW was positioned at the bedside, 0.85m from the aerosol source for all conditions. **B)** For experiment 2, the HCW was positioned either at the same beside position (0.85m) or at a distanced location (2.70m) from the nebuliser. In experiment 2 the HEPA filter was positioned at the foot and opposite side of the bed to the HCW, 2.15m from the nebuliser.

A HCW wearing PPE remained seated in the room during nebulisation (∼40mins), in one of two locations (see Figure 1). The ‘bedside’ location was positioned 0.85m from the nebulizer, whereas the ‘distanced’ location was at 2.70m.

After exiting the room, the HCW doffed PPE according to a standardised protocol (see supplement methods). Skin/nasal swabs were used to quantify HCW contamination from viruses infiltrating PPE during the exposure period. All swab samples were collected by a single experimenter (SL). Swabs were immersed in 3mL of 1X PBS in a test tube, and applied individually to 5 separate areas: forearms, neck, forehead, mouth/nose under the mask, and inside nostrils (swabbed 360 degrees 1-2cm within the nasal vestibule), see supplemental Figure S2. After application, swabs were re-immersed in PBS and sealed within the test tube. One mL of PBS (with swab immersed) was collected and plated neat with bacterial host to obtain a plaque count from each swab. After swabs were collected the HCW then showered according to a standardised protocol (see supplementary Table S1) and skin swabs were then repeated post-shower.

The settle plate method was used quantify environmental contamination from virus aerosol^18,19^. Settle plates were prepared by adding 1 mL of an overnight culture of E coli C to a lysogeny broth agar plate using the soft agar overlay method. Thirteen settle plates were positioned and left uncovered during each nebulisation (see Figure 1). After nebulisation, plates were sealed, incubated overnight at 37°C and viral plaques were enumerated the following day. Quantification of plaque counts (from swabs and settle plates) was performed by a single experimenter who was blinded to experimental conditions (DS).

### Experimental protocols

#### Experiment 1

To assess the efficacy of PPE to protect against virus aerosol exposure, the HCW was seated at the bedside during nebulisation. Virus counts from face and nasal swabs were used to assess the relative efficacy of the: 1) surgical mask, 2) poor fitting N95 mask and 3) fit-tested N95 mask. A gown, gloves and face shield were worn in each condition. A condition in which no PPE was worn was used as a positive control. Each mask condition was replicated 5 times across 5 testing days on which conditions were performed in a randomised order.

#### Experiment 2

To assess the efficacy of combining multiple control measures. The same experimental paradigm was used with constant HEPA. The HCW was seated either bedside (0.85m from aerosol source) or at a distanced location (2.70m from the source), with the HEPA filter placed at the foot of the bed (see Figure 1). The HCW wore either a surgical mask or fitted N95 mask. A gown, gloves and face shield were worn in each condition. Each condition was replicated 3 times (condition order randomised).

### Personal Protective Equipment

All experiments were performed on a single HCW (SJ) who wore a gown (Virafree Isolation Gown, Jiangxi Fashionwind Apparel Co. Ltd.), disposable gloves (Nisense nitrile gloves, Mediflex Industries) and faceshield (PET Face Shield, Xamen Sanmiss Bags Co).

The efficacy of 3 mask variants were tested:

- 3-ply surgical mask (Premium face mask, OBE Care, Singapore)
- Poor fitting (fit-factor<100) N95 respirator (N95 Healthcare Particulate Respirator, BYD Precision Manufacture Co., Ltd.)
- Fit-tested (fit-factor=194) N95 respirator (3M Aura 9320A+, 3M Australia)

Quantitative fit testing was performed via TSI PortaCount Fit Tester Model 8048. Masks were individually fit checked by the wearer on each application to optimise fit. Breathing was at rest and was predominantly nasal with periods of oral breathing to check/confirm mask fit. The HCW was clean shaven prior to each experiment to reduce mask/beard interactions. After aerosol exposure, PPE was doffed in a room separated from the clinical room by a corridor and 4 sealed doors. The doffing room had continuous HEPA filtration (5 exchanges per hour). The doffing procedure was videoed and examined independently by two expert nurses (SD, DK) to ensure doffing procedure compliance (see supplementary methods).

### HEPA filtration

For all experiments the IQAir HealthPro250 was used at its highest filtration rate 470m^3^/hr (13.4 filtration volume exchanges based on room volume). This device was also run for 30 minutes (approximately 6.7 filtration exchanges) after nebulisation was completed to purge the room of bacteriophages before repeating experimental conditions. Purging of the room was confirmed by deployment of control plates.

### Data analysis

In each experiment viable viruses were quantified by counting the number of viruses from swabs and on settling plates. Virus counts >200 were considered too-many-to-count (TMTC) and rated using an ordinal visual rating scale (+, ++, +++, ++++), with TMTC++++ indicating complete lysis of the plate’s bacterial host.For graphing/analysis, TMTC ratings were given values of 200, 210, 220, and 230. Wilcoxon, Mann Whitney U, or Friedman’s test with post-hoc comparisons (uncorrected Dunn test) was used to compare virus counts between conditions.

## RESULTS

### Experiment 1: The efficacy of PPE to mitigate MCW exposure from virus aerosol

Settle plates confirmed substantial virus contamination of surfaces in the clinical room across all experimental conditions (see supplementary Figure S3).

Virus counts recovered from swabs of each skin site are shown in Figure 2. Virus counts recovered from skin under the mask significantly differed by mask type (χ_Friedmans_=9.08 p=0.028, Figure 2A), however only the fit-tested N95 mask resulted in significantly lower virus counts compared to the no mask control (p=0.027). Given the very high and variable virus counts recovered from the skin under the mask, skin swabs taken from within the nostril were introduced on the 3^rd^ experiment day (i.e. 3 repetitions are available). Virus counts from inside the nostril (Figure 2B) were consistently high for control, surgical and poor-fitted N95 mask conditions. There was a trend for the fit-tested N95 mask to reduce virus counts on nasal swab (p=0.058), however positive virus counts were still recovered on all tests.

**Figure 2.**
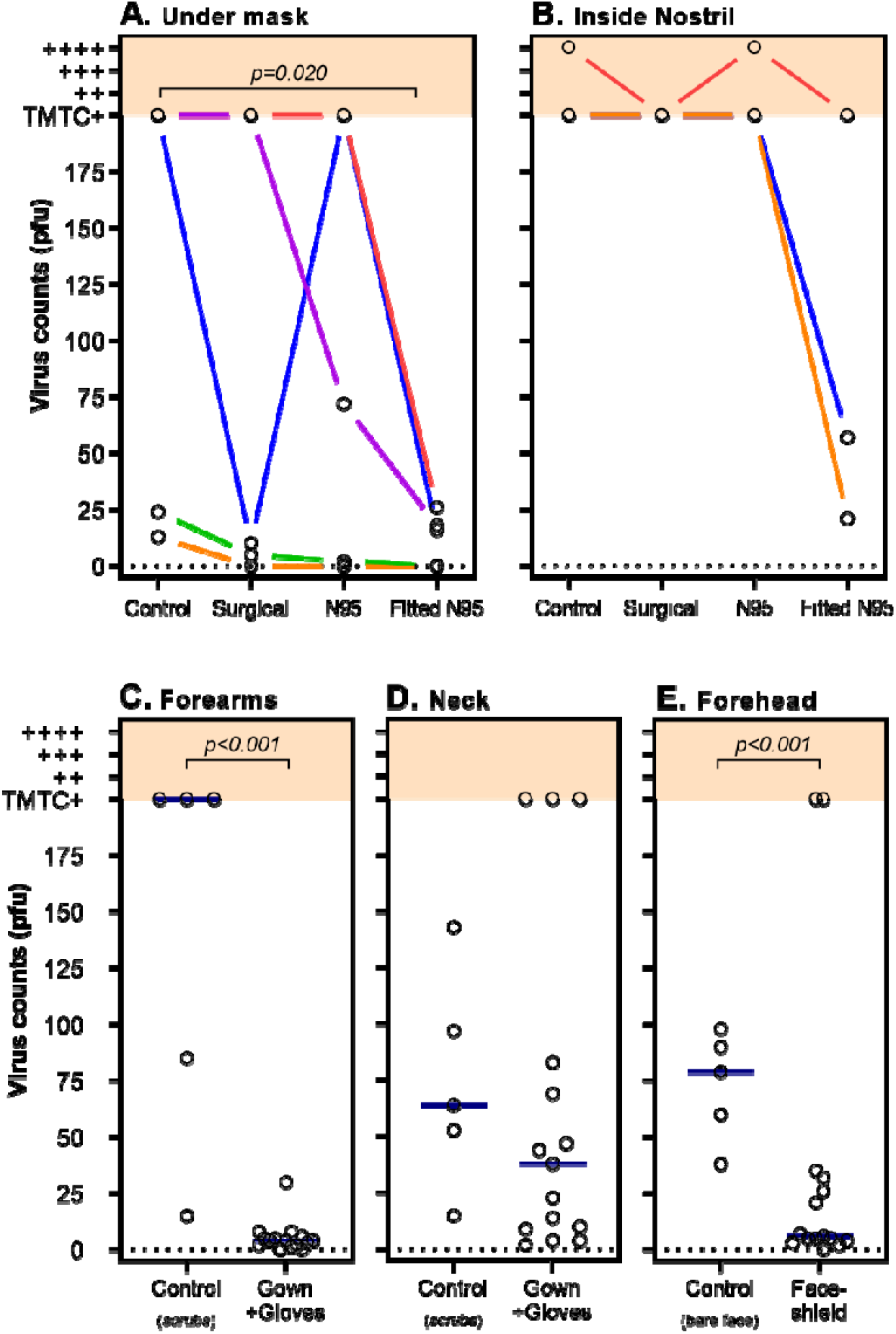
The effect of PPE on Virus plaque counts. Virus counts recovered from skin swabs (open circles, y-axis) are shown and the mitigating effect of differing types of PPE (conditions described on x-axes). Virus counts were quantified as plaque forming units (PFU) as previously described^16^. Virus counts >200 were considered too-many-to-count (TMTC) and were rated using an ordinal (+, ++, +++, ++++, shown in orange shading) visual rating scale. Blue bars represent median values. **A)**. Virus counts measured around mouth/nose underneath mask. Compared to the non-masked control condition, virus counts were found to be significantly lower when a fitted N95 mask was worn (p=0.0174). Coloured lines represent data collected on same day (in randomised order). While there is distinct variability in virus counts within conditions, data collected on same day (with same bacteriophage titre) show consistent trends of reduced virus counts for the fitted N95 mask. **B)**. Virus counts were the highest when measured from inside the nostril. There was a trend (p=0.058) for a fit-tested N95 mask to reduce virus counts. However, a surgical mask and poor fitted N95 did not appear to mitigate virus exposure. **C)** Virus counts were substantially lower on forearms/back of the hands when a gown and gloves was worn compared to a control condition in which no PPE (only scrubs) was worn (p<0.001). **D)** Virus counts on the neck were not significantly reduced by a gown with an exposed neck, compared to no PPE control condition (p=0.297). **E)** Virus counts recovered from forehead swabs were significantly lower when wearing a face shield compared to the no PPE control condition (p=0.014).

As shown in Figure 2 there were highly variable virus counts within conditions, particularly under the mask. This variability was most notable between testing days, most likely driven by differences is the bacteriophage titre. Whereas within a day (in which all mask types were compared in a randomised order, coloured lines Figures 2A and 2B) there were consistent trends to suggest that the fitted N95 mask always performed superior to the control condition. Similarly, surgical and N95 masks were largely superior (in all but one case) when compared to control.

To assess the efficacy of gloves, gown, and face shields to reduce virus counts recovered from the body, data were combined across all mask conditions and compared to the no PPE control condition (see Figures 2C, 2D & 2E). A gown combined with gloves substantially reduced virus counts on the forearms/back of hands compared to the no PPE control (U=1, p<0.001). Viruses were detectable on all neck samples and there no significant difference between the no PPE control and when the gown was worn (noting the gown used does not cover the neck). Virus counts measured from forehead swabs were significantly reduced when the faceshield was worn compared to the no PPE control (U=1.0, p=0.014).

### Experiment 2: The efficacy of combining PPE, HEPA filtration and distancing

Settle plates showed substantial virus counts during nebulisation despite the presence of the HEPA filter. Settle plates closest to the aerosol source (Plates 4 and 5, see Figure 1) demonstrated the highest virus counts. Overall, virus counts from plates were significantly lower with HEPA filtration compared to virus counts on plates from Experiment 1 (supplementary Figure S5).

As shown in Figure 3, virus counts were on average lower across swab locations and conditions compared to Experiment 1. Virus counts from under the mask differed between conditions (χ_Friedmans_=7.93 p=0.028, Figure 3A) and were higher at bedside with a surgical mask compared to the fitted N95 mask (p=0.027). There was trend suggesting the fitted N95 mask at bedside outperformed the surgical mask at distance (p=0.058). Most notably virus counts were near zero for the fitted N95 mask at both distances.

**Figure 3.**
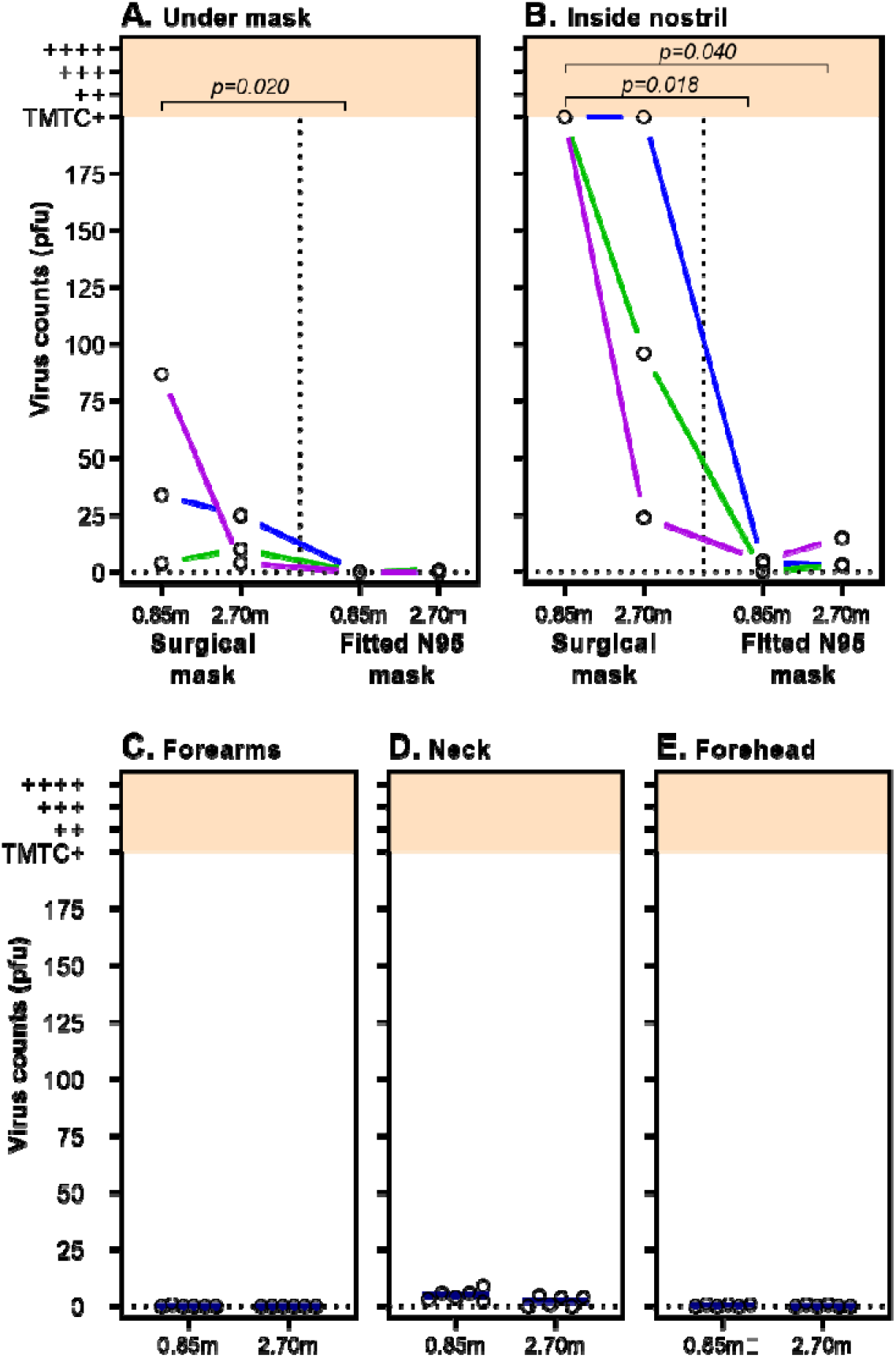
HEPA filtration combined with PPE and distance on virus plaque counts. Virus counts from skin swabs (open circles, y-axis) are shown at 0.85m (i.e. bedside) and 2.5m (distanced) locations (x-axis). Virus counts were quantified as plaque forming units (PFU). Coloured lines connect data points collected on the same day (same exact bacteriophage titre). Blue bars represent the median. A HEPA filter set to a clean air filtration rate of 470m^3^/hr (equivalent to 13 exchanges/hr) is present in all conditions. **A)** Virus counts recovered from under the mask were significantly lower with a fitted N95 mask. **B)** Virus counts from inside nostril were substantially higher for the surgical mask compared to the fitted N95 mask. Combining HEPA filtration and PPE (gown, gloves and face shield) resulted in very low virus counts on **C)** Forearms, **D)** Neck, and **E)** Forehead, however the neck was the least protected body site, likely due to no coverage provided by the gown.

Consistent with Experiment 1, virus counts recovered from nasal swabs were on average higher than all other swab sites and varied substantially via mask type and distance (χ_Friedmans_=7.97, p=0.017, Figure 3B). Nasal virus counts under a surgical mask were consistently too-many-to-count and significantly higher than the fitted N95 mask at both bedside (p=0.017) and distanced (p=0.04) positions. While nasal swab virus counts were lower with the N95 at bedside compared to the surgical mask at distance, this was not statistically significant (p=0.08), similar to the results for swabs under the mask. Virus counts did not significantly differ based on distance for surgical masks (p=0.52), or for the fitted N95 mask (p=0.75, likely to due to floor effect).

The combination of PPE protecting the body (gown, gloves, face shield) and HEPA filtration resulted in very low virus counts on the forearms and forehead, regardless of distance from aerosol source. The highest virus count recovered from neck or hand swabs in all experiments was 1 PFU. The neck site had low virus counts (median [IQR], bedside: 5 [2.75 to 7.75], distanced: 2.5 [0 to 4.25]), but were consistently higher compared to forehead swabs. The difference between bedside and distanced was not statistically significant (p>0.99).

### The effect of showering on virus counts

To quantify the effect of showering to reduce virus counts, swab data from both Experiment 1 and 2 were combined (regardless of PPE/distance/HEPA condition). Virus swab counts were compared between the post exposure swabs and after shower swabs, for each swab position.

As shown in Figure 4, virus counts were substantially reduced to near zero levels following a shower. The reduction in virus counts was statistically significant for each swab site. A supplementary analysis comparing the effect of shower pre-post the no PPE control conditions is shown in Figure S6. These data similarly show strong reductions in virus counts to near zero levels despite high post-exposure swab virus counts.

**Figure 4.**
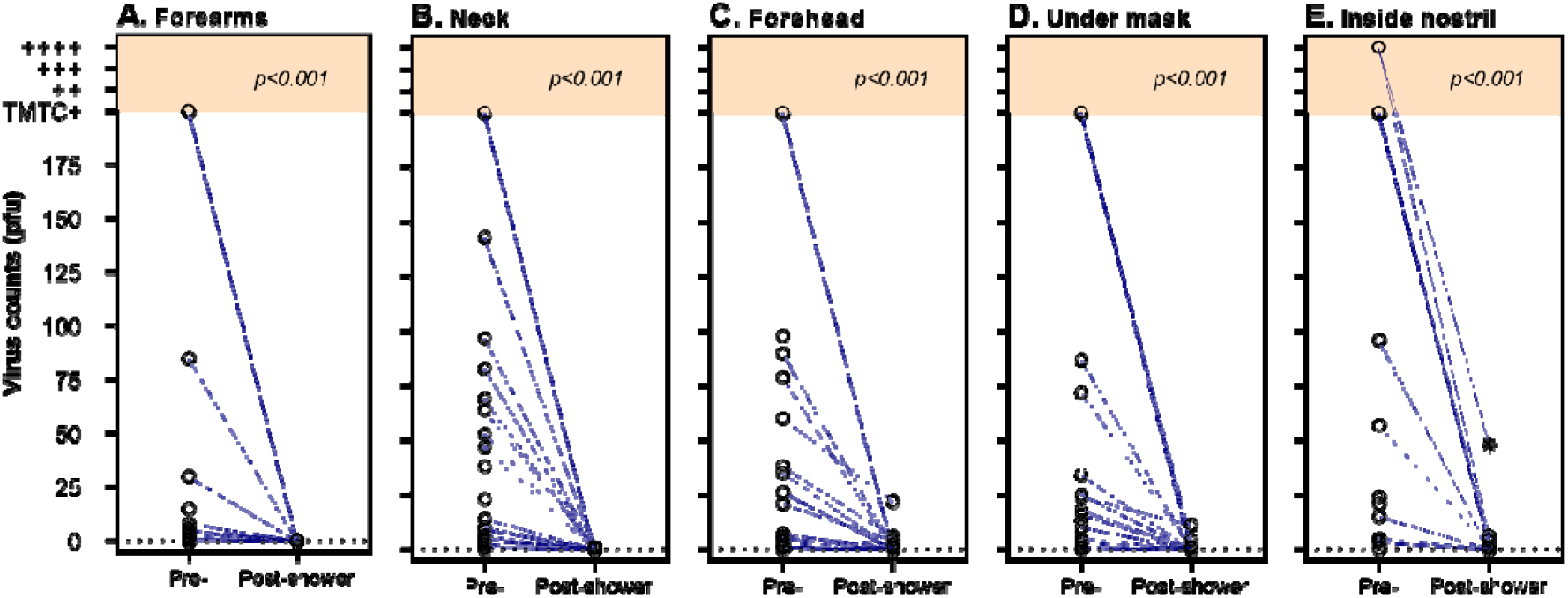
Virus counts pre- and post-shower. Virus counts recovered from skin swabs (open circles, y-axis) are shown pre-shower (i.e. post virus aerosol exposure) and post-shower (x-axes). Virus counts were quantified as plaque forming units (PFU). Virus counts are combined across all PPE conditions from experiments 1 and 2. Virus counts from **A) Forearm/hands** (median [IQR]; 3 [0 to 8] vs 0 [0 to 0] PFU, W=-120, p<0.001), **B) Neck** (9 [4 to 76] vs 0 [0 to 0] PFU, W=-276, p<0.001), **C) Forehead** (5 [1 to 36.5] vs 0 [0 to 1] PFU, W=-224, p<0.001), **D) Under mask** (16 [0 to 143.5] vs 0 [0 to 0] PFU; W=-187, p<0.001), and **E) Inside nostril** (TMTC [12.5 to TMTC] vs 0 [0 to 1.75] PFU, W=-105, p<0.001) swabs were all significantly reduced to near zero levels following a shower. There was one post shower nasal swab that resulted in a high virus count of 48 PFU (shown as a star), this corresponded to first nasal swab sample collected. Due to this high residual virus count, the shower protocol was changed to include a nasal rinse with a saline solution.

## DISCUSSION

Our study is the first to use live virus aerosol experiments to systematically examine HCW virus contamination and the interaction between virus aerosol, PPE and air filtration using a portable HEPA filter. The most important finding of our study is that the combination of a quantitatively fit tested N95 mask and portable HEPA filter (filtering the total room volume 13.7 times per hour) provides near complete protection against extremely high virus aerosol loads (10^9^ copies) at close range (0.85m) for prolonged periods of time (45mins). Critically, surgical masks provided inadequate protection against skin and upper airway contamination, even when combined with HEPA filtration and at distances of 2.70m. We also found that a gown, gloves, and face shield do not protect against skin contamination of the neck and forehead, but that showering according to protocol can almost completely remove virus contamination. In light of aerosol transmission of SARS-CoV-2 and the emergence of more transmissible variants of concern, our findings have immediate and broad implications for the protection of HCWs.

Clinical evidence for the superiority of N95 respirators over other mask types in protecting health care workers against SARS-CoV-2 infection is mixed^20–22^. Our data elucidates two possible mechanistic reasons for mixed clinical signals: 1) mask fit and 2) fallibility at high viral load. Approved N95 respirators perform to a filtration standard that protects against particles to the nanometre range^23^. Importantly, previous reports of surgical and N95 mask penetration properties show that peripheral leak is more important than the filtering properties of the mask material^24^. Gaps between the face and mask provide low resistance points for airflow to circumnavigate the (higher resistance) mask filter. Poorly fitting masks therefore allow significant airflow through these gaps into which virus laden aerosol can infiltrate. Our study demonstrates that a quantitatively fit-tested N95 mask reduces skin and nasal virus aerosol contamination compared to poor fitting N95 and surgical masks. Importantly, both the poor fitting N95 and surgical masks were fit checked at time of each application to ensure the best possible fit for that specific mask during each condition. Fit tested N95 was the only condition that proved superior to control (no mask). The poor fitting N95 performed with similar efficacy as surgical mask, again highlighting the critical importance of mask fit. It reinforces the necessity of quantitative fit testing of N95 respirators for all forward-facing HCWs, a process that is not universal practice and that relies on available mask supply^25^. However, even with the best fitting N95 mask there was still significant virus aerosol contamination of the nose after 45mins exposure to high virus aerosol load (10^9^ copies) at close range (0.85m) in the absence of HEPA filtration (see Figure 2, panel B).

In our study we demonstrated two significant effects of HEPA filtration on virus laden aerosol. Firstly, in relation to HCWs, we found that a HEPA filter (providing 13.7 volume filtrations/hour) enhances the PPE effectiveness so that a quantitatively fit-tested N95 mask provided almost complete protection against skin and nasal contamination from virus aerosol. The level of protection offered from nasal virus deposition is critical because it relates directly to deposition of virus in the lungs – the primary site of SARS-CoV-2 entry. Although particle deposition in the respiratory tract is complex, we can extrapolate from previous studies the likely relationship between nasal and lung deposition of virus particles in our study. An extensively verified model for regional aerosol deposition shows that for molecules in the range 0.2-10µm, deposition in the anterior nose is greater than that in the distal bronchial tree (bronchioles) when breathing through either the mouth or nose^26^. This is confirmed by in vivo gamma scintigraphic studies utilising continuous Pari-PEP nebulisation with exclusively oral breathing, which show that lung deposition of aerosol of mass median diameter 4µm is less than nasal deposition (14.2% upper airway vs 12.8% whole lung deposition)^27^. As such, the finding in our study of almost complete protection from virus aerosol on nasal swab can be used to infer almost complete protection from lung deposition. This is critically important for the protection of HCWs in the COVID-19 context. Secondly, in relation to hospitalised patients – we found that HEPA filter deployment reduced virus counts on room settling plates compared to the no HEPA filter condition. However, there was still extensive environmental virus contamination with the HEPA filter deployed. This has clear implications for the deployment of such devices in hospital environments. Although the HEPA filter reduced aerosol load and therefore likely reduces risk, it does not completely negate environmental contamination in the same way as a point of emission strategy does^18^.

Our study demonstrates that areas of exposed skin such as the forehead and neck become contaminated with virus aerosol when PPE is deployed without the addition of a HEPA filter (including contamination of nose, chin and cheeks under the mask). Previous studies have demonstrated that over a 4 hour period, people touch their chin/cheek 792 times, and neck 104 times^28^. This apparently innate human behaviour can transfer virus from skin to mucous membranes (eyes, nose, mouth), which people touch with their hands 15.7 times every 3 hrs^29^. Our study demonstrates that showering according to protocol almost completely removes virus from the face, neck, and forehead. While there are some instances of low virus counts detectable after showering (see Figure 4), PhiX174 is far more robust to inactivation by soap compared to lipid enveloped respiratory viruses such as SAR-COV-2 and influenza. Therefore, showering would be expected to have greater efficacy at protecting HCWs against such viruses. Importantly, a Cochrane review of barriers to healthcare worker adherence to infection control practice, identified a lack of showering facilities as a major barrier^30^. The importance of showering is clear, but any delay between doffing PPE and showering increases the risk of inadvertent transfer of virus to mucous membranes. Workplace facilities that enable rapid shower decontamination prior to workers leaving are essential in reducing transmission risk.

There are several limitations of this study. First, we aerosolised a higher viral load (10^8^/mL) compared to viral load of aerosols generated by patients infected with coronaviruses^31^. However, the choice of bacteriophage titre was determined based on detection sensitivity experiments (for skin swab and settle plates), was similar to previous experiments by our group^18^ and others^32^ and allows us to quantify degree of contamination in a way that relates directly to the viral load titre and detection sensitivity. Most importantly, the use of high viral load provides an appropriately strong safety test of these PPE strategies. Our method has clear advantages over smoke/chemically generated aerosols, as it can demonstrate contamination/infiltration with “live” virus on skin, surfaces, and under PPE. Second, we observed significant variability in viruses quantified from swab samples. Our analyses demonstrate that much of this variability is driven by between day differences, likely resulting from differences in the bacteriophage titre aerosolised on a given day. Our within-day data demonstrate consistent relative reductions in virus counts between the fitted N95 mask and the control (see Figures 1A and B). This is important given that a high degree of variability in the viral load/exposure is an expected phenomenon in health care settings. Although a single experimenter conducted all swabbing procedures, the performance/accuracy of skin swabs are likely to be affected by the pressure with which the swab is applied, and the rotation swab. Once the swab is returned to the laboratory it is assumed all virions contained in the swab tip do not necessarily dissipate into the phosphate buffered solution for quantification. The advantage of using the swab technique is that it can be applied to defined areas multiple times (i.e. repeated per condition) and it is easy to perform and cheap to deploy. Importantly, for our main finding of the effect of quantitative fit tested N95 and HEPA filter, the results indicating almost universal absolute zero are reassuring in this regard. Third, our experiments are conducted in a sealed clinical room. Due to this, no mixing between clean/external air is occurring (e.g. ingress under door gap or via ventilation openings). As such we feel the lack of mixing biases towards a higher aerosol load in the experimental room. However, this again provides reassurance about the high effectiveness of the combination of fitted N95, PPE and HEPA filter in reducing HCW virus contamination to near zero. Finally, although particle analysis was not an endpoint of our study, indicative reference recordings in our experiments demonstrate a high particle load of size distribution in the range of that generated by humans^33^.

In conclusion, the emergence of more transmissible variants of SARS-CoV-2 have highlighted the gaps in protecting HCWs that were exposed in 2020. Health care providers must deploy a simultaneous array of mitigation strategies to optimise HCW safety. We have demonstrated that quantitatively fit tested N95 masks combined with a HEPA filter can offer protection against high virus aerosol loads at close range, for prolonged periods of time. Any skin contamination that occurs in the workplace can be removed by showering according to protocol.

## Data Availability

Data is available online, link is in the supplement.

