## Supplementry Materials for "Personal protective equipment does not sufficiently protect against virus aerosol unless combined with advanced air purification or ventilation techniques"

Shane A Landry^1^ PhD

Dinesh Subedi^2^ PhD

Jeremy J Barr^2^ PhD

Martin I MacDonald^3^ PhD

Samantha Dix^4^

Donna M Kutey^4^

Darren Mansfield^3,5,6^ PhD

Garun S Hamilton^3,5,6^ PhD

Bradley A. Edwards^1,2^ PhD

Simon A Joosten^3,5,6^ PhD

**Affiliations**

1. *Department of Physiology, School of Biomedical Sciences & Biomedical Discovery Institute, Monash University, Melbourne, VIC, Australia*
2. *School of Biological Sciences, Monash University, Clayton, VIC, Australia*
3. *Monash Lung, Sleep, Allergy and Immunology, Monash Health, Clayton, VIC, Australia*
4. *Monash Nursing & Midwifery, Monash University, Clayton, VIC, Australia*
5. *School of Clinical Sciences, Monash University, Melbourne, VIC, Australia*
6. *Monash Partners – Epworth, Victoria, VIC, Australia*
7. *Turner Institute for Brain and Mental Health, Monash University, Melbourne, VIC, Australia*

**Correspondence:**

Author:	Shane Landry, PhD

Address:	Sleep and Circadian Medicine Laboratory

Ground Floor, Monash University BASE facility

264 Ferntree Gully Road

Notting Hill, 3168, Victoria, Australia

**Supplementary Methods**

*Clinical room*

The same clinical room was used in all experiments. To control airflow patterns, ceiling vents were taped shut and heating and cooling appliances were switched off. The room was well insulated such that temperature (median [min – max]; 23.3 [20.8 – 26.6] ºC), humidity (42 [32 – 54] %) and barometric pressure (993.8 [999.3 – 1018] mmHg) were well controlled during experimental procedures. The room door is a double sealed soundproofed/insulated door.

*Nebulisation*

The nebulizer (PARI Respiratory Equipment, VA, USA) was positioned at the bed head. A 90º plastic connector (ID=19mm) was attached the nebulizer so that aerosols were expelled vertically. Medical air (9 L/min) was delivered to the nebulizer via standard oxygen tubing. The tubing was fed through a sealed port wall port and connected to a wall mounted flow metre (RTM3 0-15 L/m) which was operated by an experimenter outside of the clinical room.

The Pari-PEP nebulizer produces a distribution of aerosol particle size of 3.42±0.15 µm^18^. We recorded particle mass concentration with a PurpleAir PA-II-SD (PurpleAir Inc. Utah, USA) sensor for reference (see Figure S1)

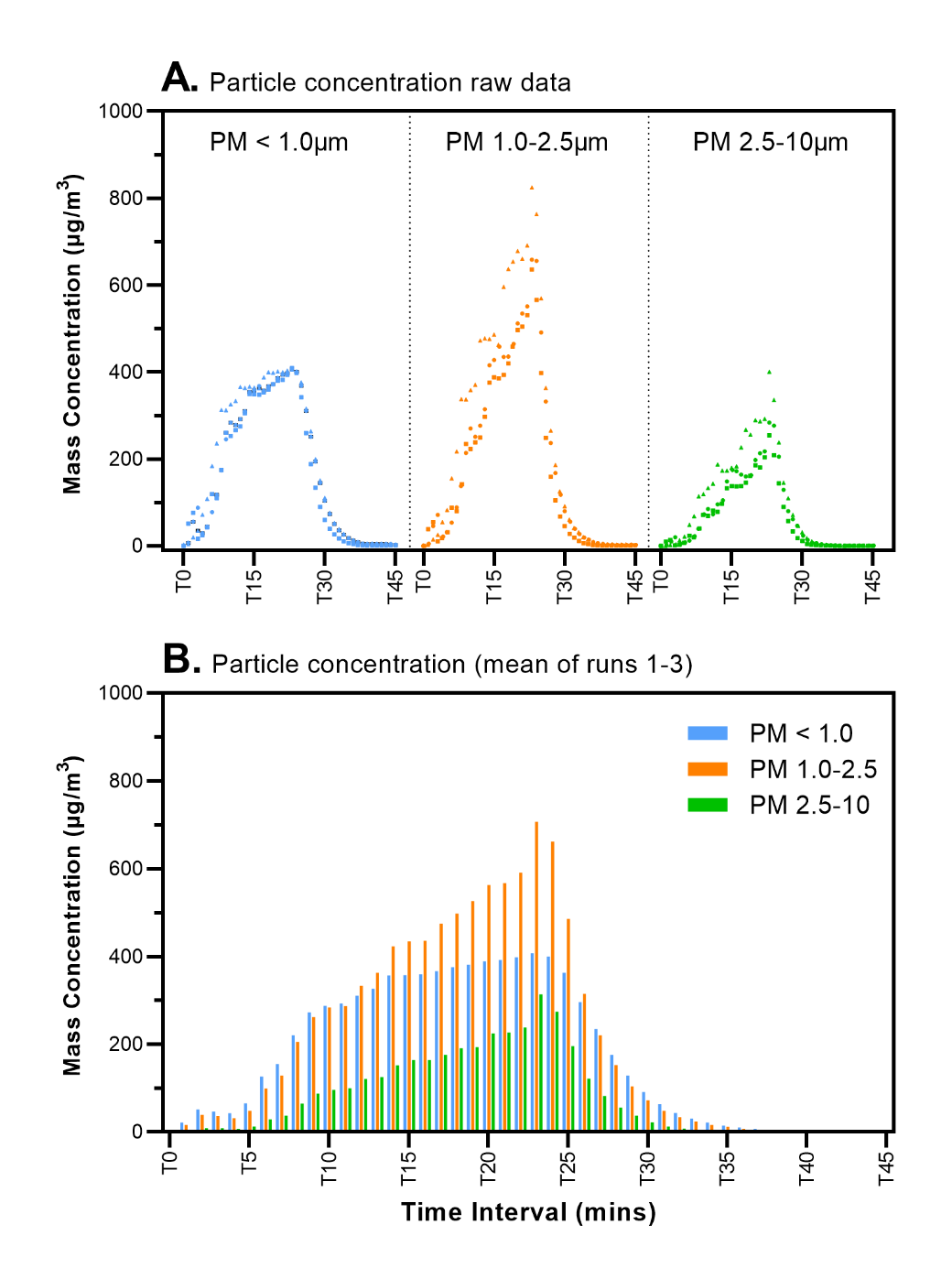

***Figure S1. Particle mass concentration during phage lysate (10mL) nebulisation with PariPEP nebuliser.*** *The particle mass concentration was measured on 3 indicative runs using a PurpleAir sensor during nebulisation of 10mL of phage lysate. Panel A) demonstrates individual raw data by time for each of 3 particle sizes. The data for each individual run is pooled by particle size. Run1=squares, Run2=circles, Run3=triangles. Panel B) demonstrates mean particle mass concentration for the 3 runs binned again by particle size. Time intervals is 2mins starting at T0. The nebuliser was switched off and the HEPA filter was switched on for room purging at during T23. PM: particulate matter.*

*Personal Protective Equipment doffing protocol*

All personal protective equipment (PPE) used in this study was donned (put on) and doffed (removed) according to local state (Victorian Department of Health and Human Services, Australia) government reference documents. These documents are published online at <https://www.dhhs.vic.gov.au/infection-prevention-control-resources-covid-19>.

Broadly, the use PPE was practiced in accordance with the COVID-19 infection prevention and control guidelines, as well as the guidelines for the conventional use of PPE.

- [COVID-19 Infection prevention and control guidelines (Word)](https://www.dhhs.vic.gov.au/covid-19-infection-control-guidelines)
- [[Guide to the conventional use of PPE (Word)](https://www.dhhs.vic.gov.au/guide-conventional-use-ppe-covid-19-doc)](https://www.dhhs.vic.gov.au/guide-conventional-use-ppe-covid-19-doc)

Specifically, the order that PPE was doffed was performed according to the following reference document:

- [Standard sequence for putting on (donning) and taking off (doffing) PPE_Jan2021 (Word)](https://www.dhhs.vic.gov.au/standard-sequence-putting-on-taking-off-ppe-covid-19-doc)

See also visual reference guide:

- [How to put on (don) and take off (doff) your PPE separately (PDF)](https://www.dhhs.vic.gov.au/how-put-and-take-your-ppe-gown-and-gloves-separately)

All doffing of PPE was video recorded. To ensure compliance with the above doffing procedures, two expert nurses with expertise in doffing procedures (DK & SD) reviewed and independently examined each video for breaches of doffing protocol. After each video was independently assessed, videos/reports were discussed to ensure concordance. After review, any breaches were rated with respect to the likelihood and severity of contamination occurring to any of the skin sites swabbed (i.e. Arms/Forearms, Neck, Forehead, Face, Nostril, see below).

These data are summarised in the online data summary document and all individual reports and videos have been uploaded to an online repository:

[<https://cloudstor.aarnet.edu.au/plus/s/igztKvpB0tMT0Q2>](https://cloudstor.aarnet.edu.au/plus/s/igztKvpB0tMT0Q2)

*Application of swabs to skin sites*

Skin surface and nasal swabs were used to quantify health care worker (HCW) contamination for experiments 1 and 2. Swabs were immersed in 3mL of 1X phosphate buffered saline contained in a test tube, and then applied individually and systematically to 5 separate areas (see Figure S1). Large size swabs (Jumbo Swabs, Multigate Medical Products Pty Ltd.) were used for all skin surface areas. For nostrils sites a smaller swab (Swisspers cotton tips, McPhersons Consumer Productions Pty Ltd) was used.

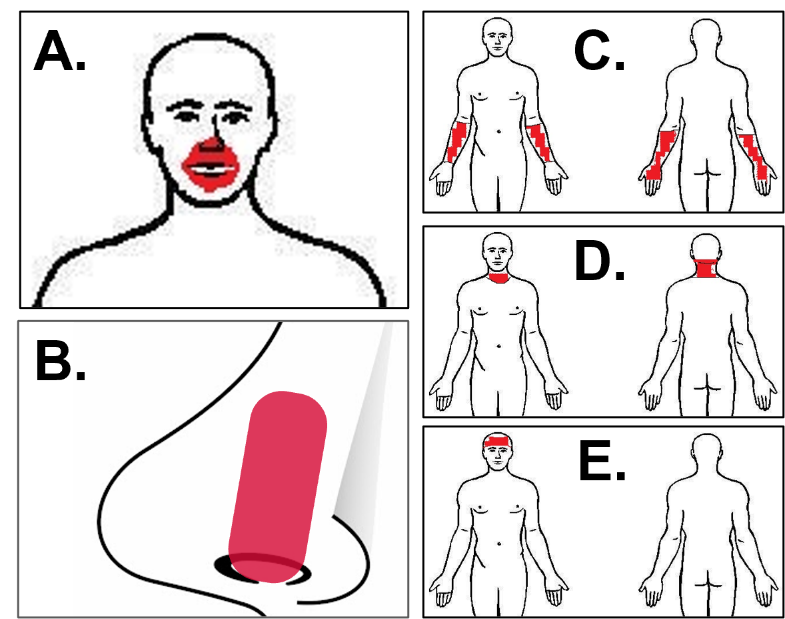

***Figure S2. Swab sites.*** *The following sites (highlighted in red) were swabbed after exposure****.***
 ***A.*** *Under the mask (around mouth/nose, specifically under mask coverage),* ***B.*** *Inside nostrils (swabbed 2x 360 degrees 1-2cm within the nasal vestibule),* ***C.*** *Forearms (top and bottom of the forearms and back of hands),* ***D.*** *Neck (mid-line in a ring around back and front), and* ***E.*** *Forehead (above eyebrows and below hairline).*

*Showering protocol*

To assess the efficacy of a shower to reduce virus contamination the HCW showered after collection of post-exposure skin swabs. Post-shower skin swabs were collected immediately after showering. The cleaning materials and the shower procedure was standardised across all experiments. These details are outlined in Table 1.

***Table S1: Shower Protocol***

Materials:

- Shower – running at standard pressure, comfortably warm water.

- Flo Sinus Wash (ENT Technologies Pty Ltd)

- salt or premix sachets to make saline water.

- Soap (Dove – Sensitive Beauty Cream Bar, Unilever)

- Shampoo (OC Naturals, Normal Balance Shampoo, Natures Organics)

- Facewash (Neutrogena, Deep Clean Facial Cleanser, Johnson and Johnson)

- Dishwashing liquid (Shine, Antibacterial Dishwashing Liquid, Woolworths)

- Towel

Procedure:

1. Post doffing procedure – implying clean hands.

2. Mix Flo Sinus wash – can use pre-purchased sachets or half a teaspoon salt with warm water.

3. Lavage into each nostril twice, clearing saline wash completely prior to next lavage.

4. Wash Flo Sinus wash bottle with detergent/dishwashing liquid.

5. Wash hospital shoes with detergent/dishwashing liquid.

6. Soap inside of nares.

7. Rinse thoroughly.

8. Wash hair, concentrate on extending soap field to neck, around, behind ears.

9. Rinse thoroughly.

10. Wash face with facewash, use generous amount, ensure soap field covers entire forehead, face including under nose and around mouth, under chin and front of neck.

11. Rinse thoroughly.

12. Wash body with soap, try to cover every part of body, include neck front and back, tops of feet.

13. Rinse thoroughly.

14. Dry off.

A video of the protocol can be found online at:

[<https://cloudstor.aarnet.edu.au/plus/s/igztKvpB0tMT0Q2>](https://cloudstor.aarnet.edu.au/plus/s/igztKvpB0tMT0Q2)

**Supplementary RESULTS AND DISCUSSION**

A data summary document containing tabulated individual data can be found online:

[<https://cloudstor.aarnet.edu.au/plus/s/igztKvpB0tMT0Q2>](https://cloudstor.aarnet.edu.au/plus/s/igztKvpB0tMT0Q2)

*Settle plates data*

Settle plates were used in all experiments to confirm the level of environmental contamination in the clinical room from virus aerosol settling on surfaces. Data from experiment 1 (nebulisation without HEPA filtration) is shown in Figure S2. There was substantial virus contamination across all plate samples in each experimental condition, with the majority of virus counts in the too-many-too-count (TMTC) range. There were high virus counts, primarily driven by higher visual TMTC ratings (χ^2^_Friedman’s_=42.16, p<0.001) in the control condition (median=TMTC++++) compared to the surgical (median= TMTC+, p<0.001), N95 (median TMTC+, p<0.001) and fitted N95 (median= TMTC++, p=0.02) mask conditions (Figure S2A). However, there was no difference between conditions with respect to the proportion of TMTC ratings versus countable plaques (χ^2^=2.45, df=3, p=0.485). Similarly, this difference between conditions was not evident when this analysis was limited to plates less than 1m from the nebuliser (i.e. proximal to where the HCW was seated during nebulisation/exposure), which were consistently TMTC++++ across conditions.

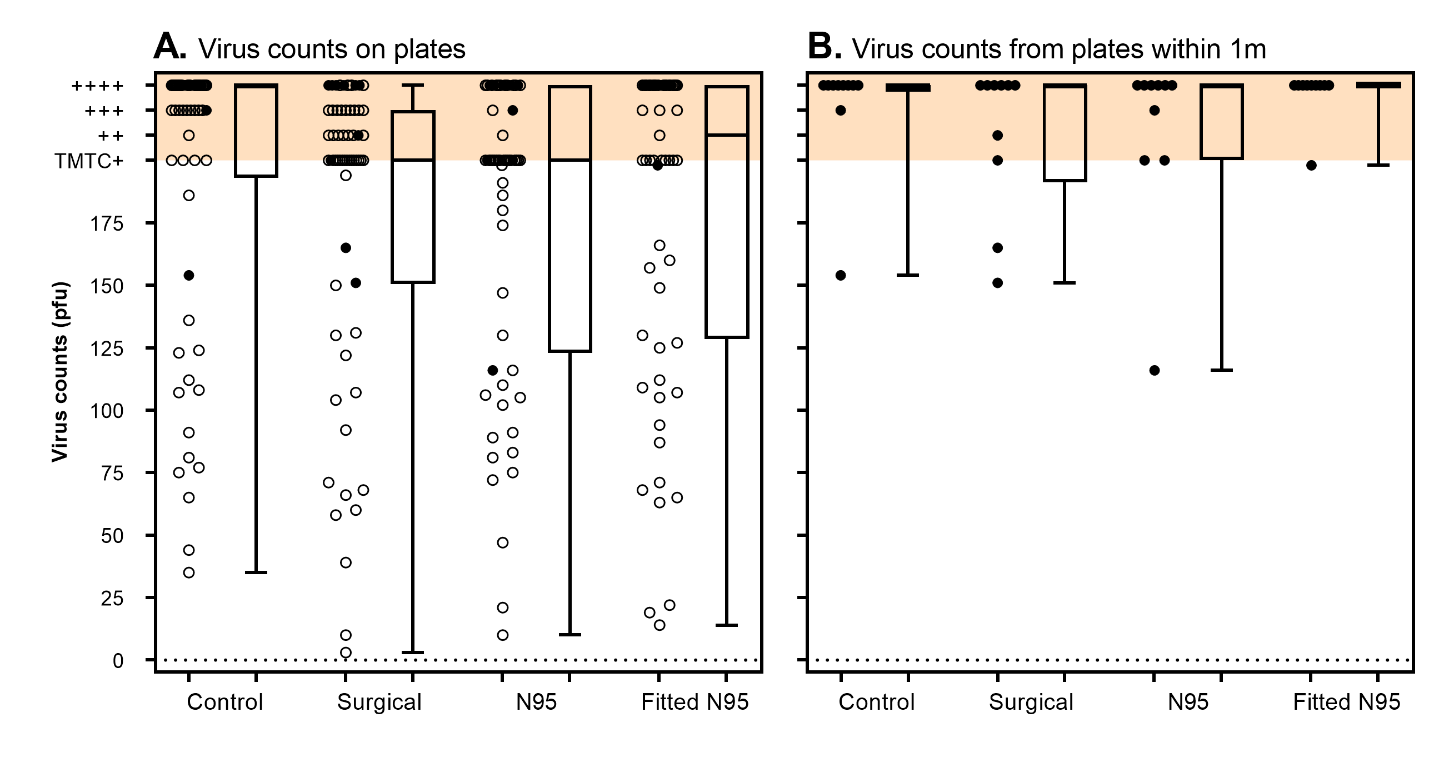

***Figure S3.*** ***Settle plate data across Experiment 1 conditions****. Virus counts from settle plates are shown for each experimental mask condition. Data are represented as individual data (circles) and as box and whisker plots. Closed circles represent virus counts from the 2 settle plates within 1m of the nebuliser (similar distance to where the HCW was seated during exposure). Virus counts were quantified as plaque forming units (PFU, y-axis). Virus counts >200 were considered too-many-to-count (TMTC) and were rated using an ordinal (+, ++, +++, ++++, shown in orange shading) visual rating scale.* ***A)*** *Shows settle plate data across all repetitions and for all 13 plates differentiated by mask conditions. Note the experimental manipulation (i.e. type of face PPE) is not expected to affect plate data).* ***B)*** *Shows virus counts on the plates specifically within 1m of the nebuliser. The HCW was seated at a similar distance for the duration of the exposure period.*

All plate data, plaque count enumerations, and TMTC ratings were assessed by a single experimenter (DS) who was blinded to the experimental conditions tested. The order of experimental mask condition was randomised across experimental days, and all 4 conditions was completed on a single experimental day to reduce the impact of day-day variation in bacteriophage titre. However, closer examination of the randomised data shows some degree of imbalance in the order. Specifically, the control condition tended to be performed earlier in the day more often (condition 1 once, condition 2 three times, condition 4 once), see Table S2.

***Table 2. Randomised condition order for Experiment 1.***

| **Order** | **16/04/2021** | **7/05/2021** | **28/05/2021** | **9/06/2021** | **11/06/2021** |
| --- | --- | --- | --- | --- | --- |
| **C1** | N95 | Control | Surgical | Fitted N95 | Fitted N95 |
| **C2** | Control | Surgical | N95 | Control | Control |
| **C3** | Fitted N95 | N95 | Fitted N95 | N95 | Surgical |
| **C4** | Surgical | Fitted N95 | Control | Surgical | N95 |

Further analysis confirms an order effect (χ^2^_Friedman’s_=12.25, p=0.007), such that settle plate exposed to virus aerosol in the first two conditions of an experimental day (C1 and C2, typically morning, medians=TMTC+++) had higher TMTC ratings compared to conditions compared to the last two conditions (C3 and C4, typically afternoon, medians=TMTC+). However, similar to the plate versus mask condition data, this difference was not significant for plates within a short distance (>1m) of the nebuliser.

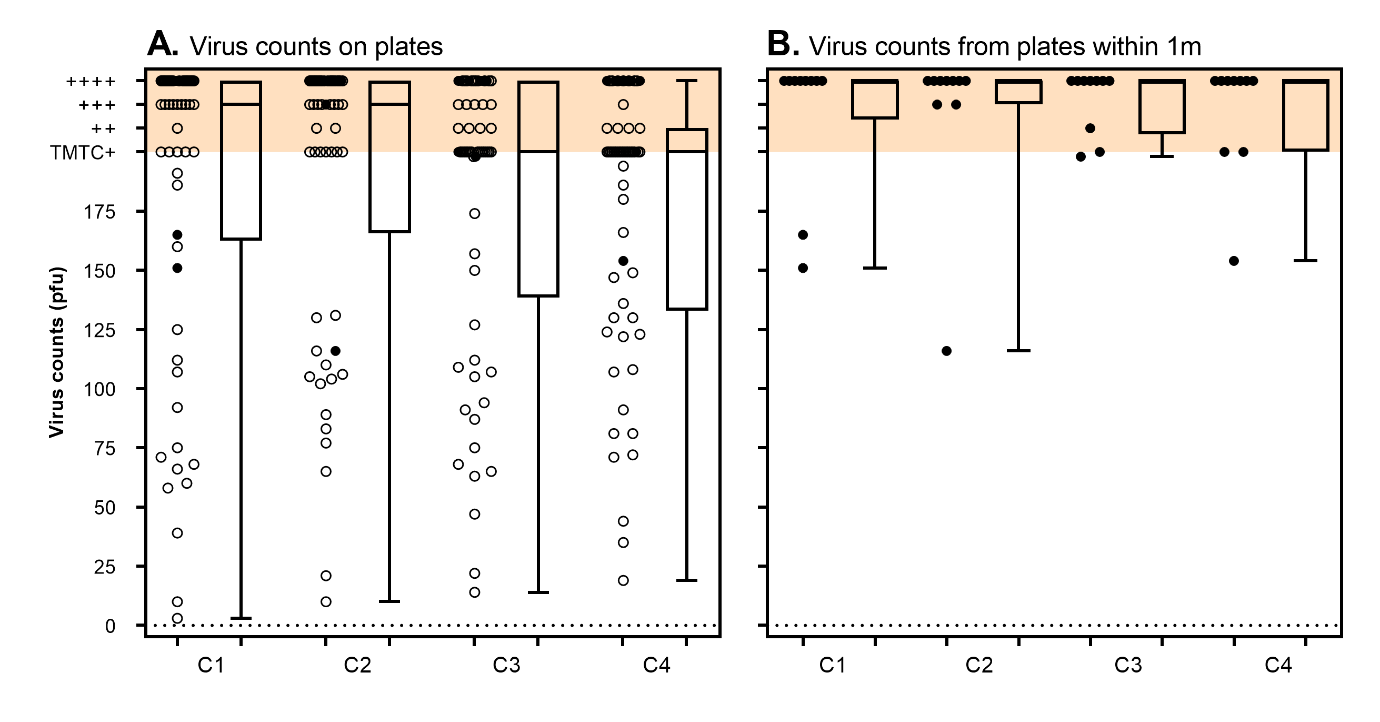

***Figure S4.*** ***Virus counts on settle plates by condition order****. Virus counts from settle plates are shown for each experimental mask condition.* ***A)*** *shows data from all settle plates.* ***B)*** *Shows data from settle plates >1m from the nebuliser.*

Given that all settle plates required for the day were prepared in the morning before each experiment, there can be minor growth of the bacterial host at room temperature over the course of an experimental day, prior to bacteriophage exposure. As a result, we observed slight changes in plaque morphology on plates used in conditions later in the day, which could have affected visual ratings (i.e. TMTC ratings) on these plates. .

| 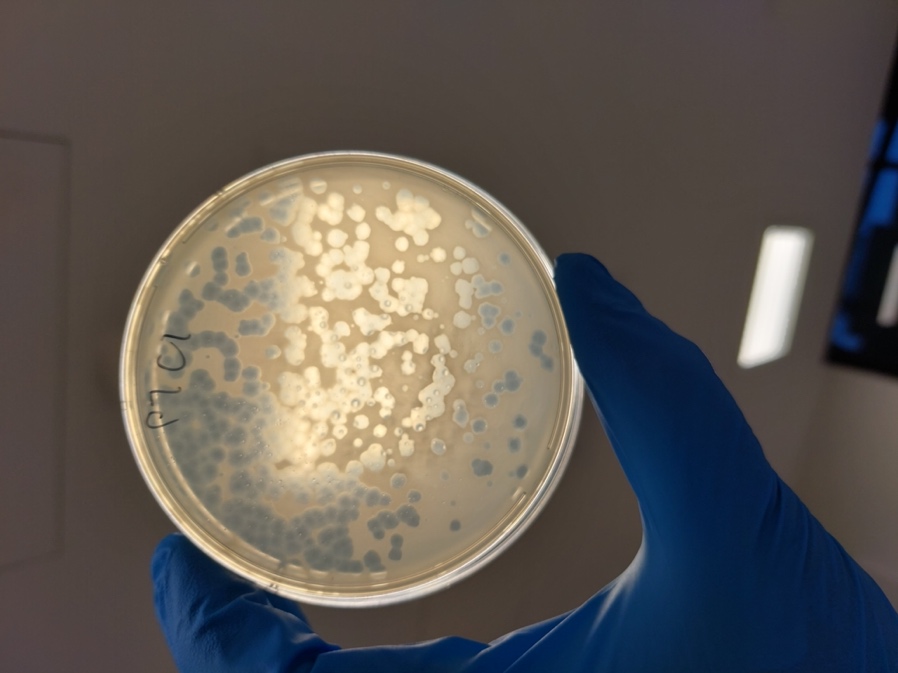 | 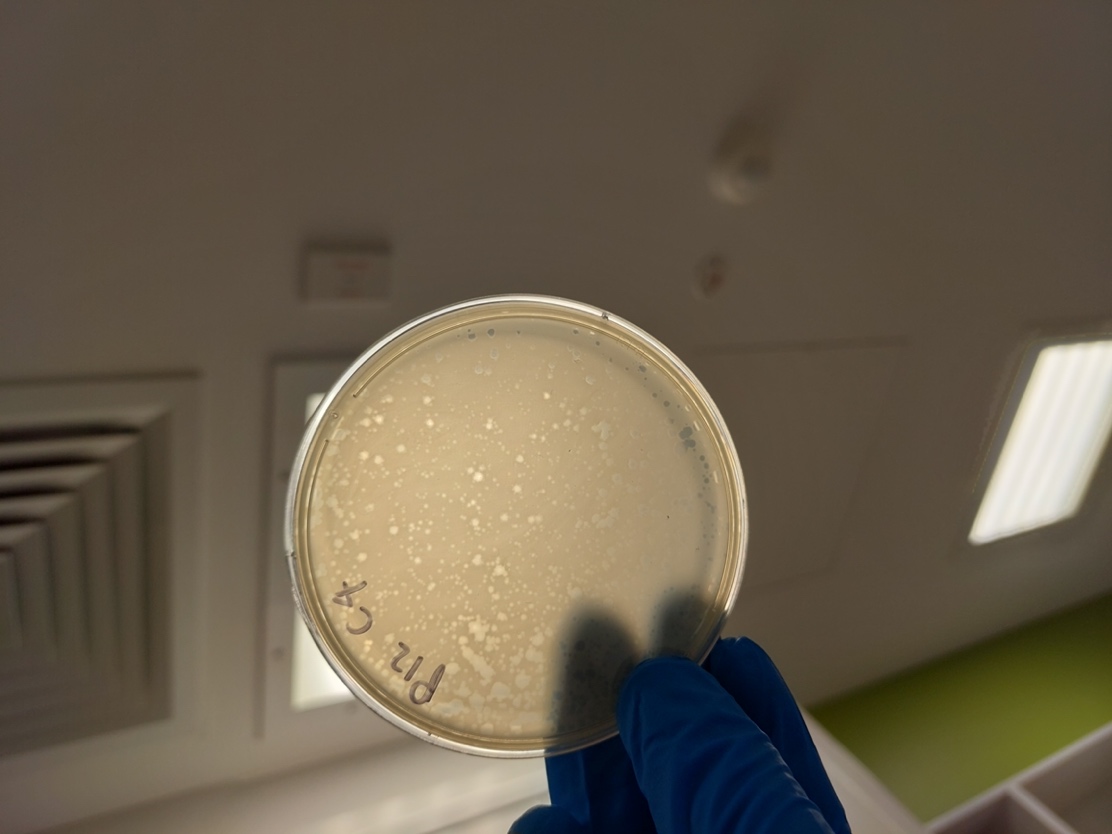 |
| --- | --- |
| *Plaques morphology on settle plates performed in the morning* | *Plaque morphology on settle plates performed later in the day* |

These data demonstrate the need to perform experiments using settle plates in a randomised order with a full Latin squares design. It should be noted that these order effects do not affect plaque counts derived from skin surface swabs (which are collected differently, i.e. swabs samples are immersed in PBS and then extracted and plated later).

*The effect of HEPA filtration on settle plates*

To explore the effect of HEPA filtration on environment contamination, all plate data from experiment 1 (without HEPA filtration) was compared to the combined settle plate data from experiment 2 (with HEPA filtration at 470m^3^/hr, ~13 exchanges). As shown in figure S4, virus counts from plates were significantly lower with HEPA filtration (median [IQR], 123.5 [65.75 to TMTC+] pfu) compared to no filtration (TMTC++ [154.8 to TMTC++++], U=10223, p<0.001). Importantly, these data show that there was still significant contamination of all plates despite air purification/HEPA filtration active throughout the entire nebulisation period. Notably there were still very high plate counts (most TMTC++++) for settle plates within 1m of the nebuliser (Figure S4B). This coincides with the bedside (0.8m) location of the HCW in experiment 1.

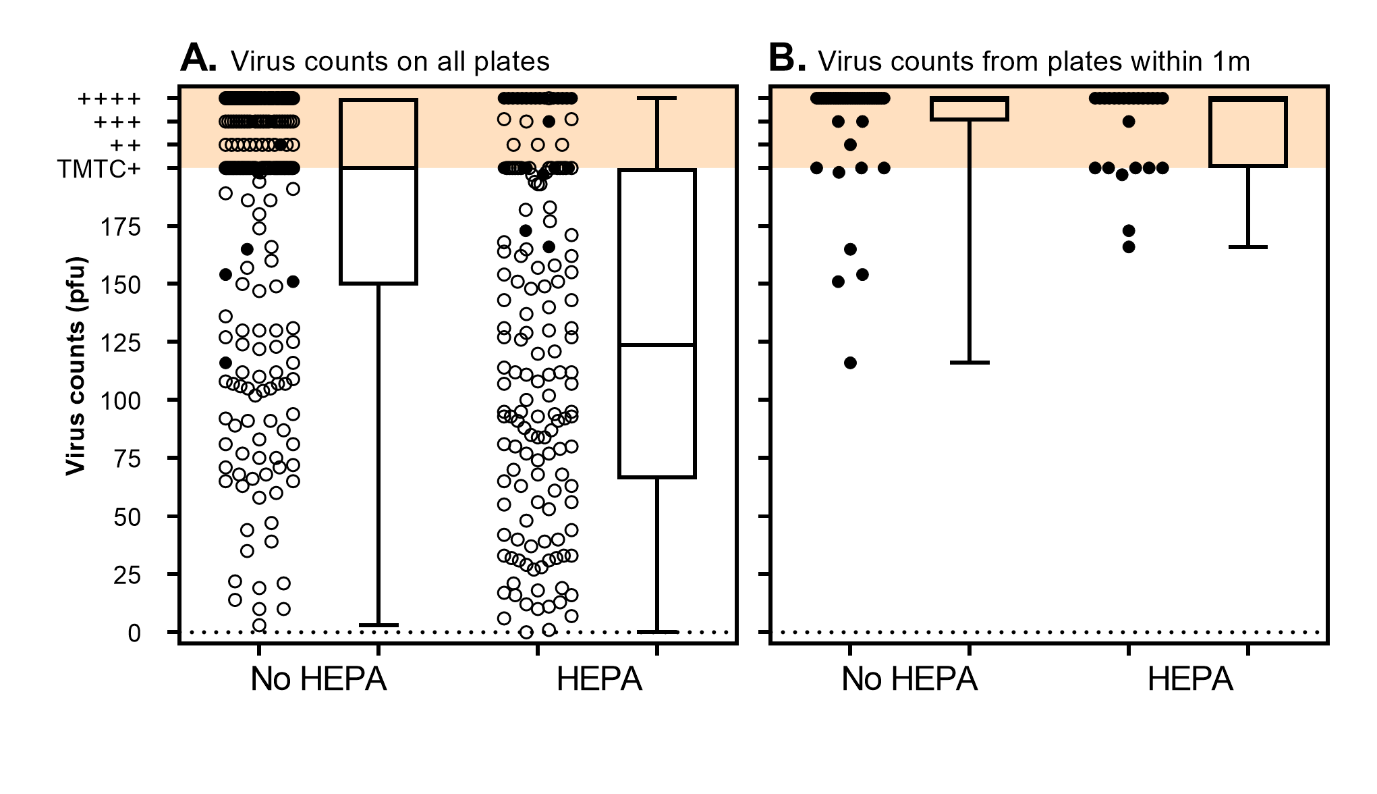

***Figure S5.*** ***The effect of HEPA filtration on plate counts****. All virus counts from settle plates are shown from experiment 1 (i.e. with no HEPA filtration) versus experiment 2 (with HEPA filtration at 470m^3^/hr, ~13 exchanges). Data are represented as individual data (circles) and as box and whisker plots. Closed circles represent virus counts from plate the 2 settle plates within 1m of the nebuliser (similar distance to HCW). Virus counts were quantified as plaque forming units (PFU, y-axis). Virus counts >200 were considered too-many-to-count (TMTC) and were rated using an ordinal (+, ++, +++, ++++, shown in orange shading) visual rating scale.* ***A)*** *shows data from all settle plates.* ***B)*** *Shows data from settle plates >1m from the nebuliser.*

*The effect of showering on virus counts*

In the main text, the effect of showering on skin swab virus counts was tested from data combined across experiment 1 and 2 (i.e. all conditions combined regardless of type of PPE/HEPA present). Here we present a supplementary analysis which compared virus counts pre-post shower, only for the no PPE control condition in experiment 1 (see Figure S5). There were large decreases in virus counts on all sites, however there were no statistically significant differences likely due to insufficient power.

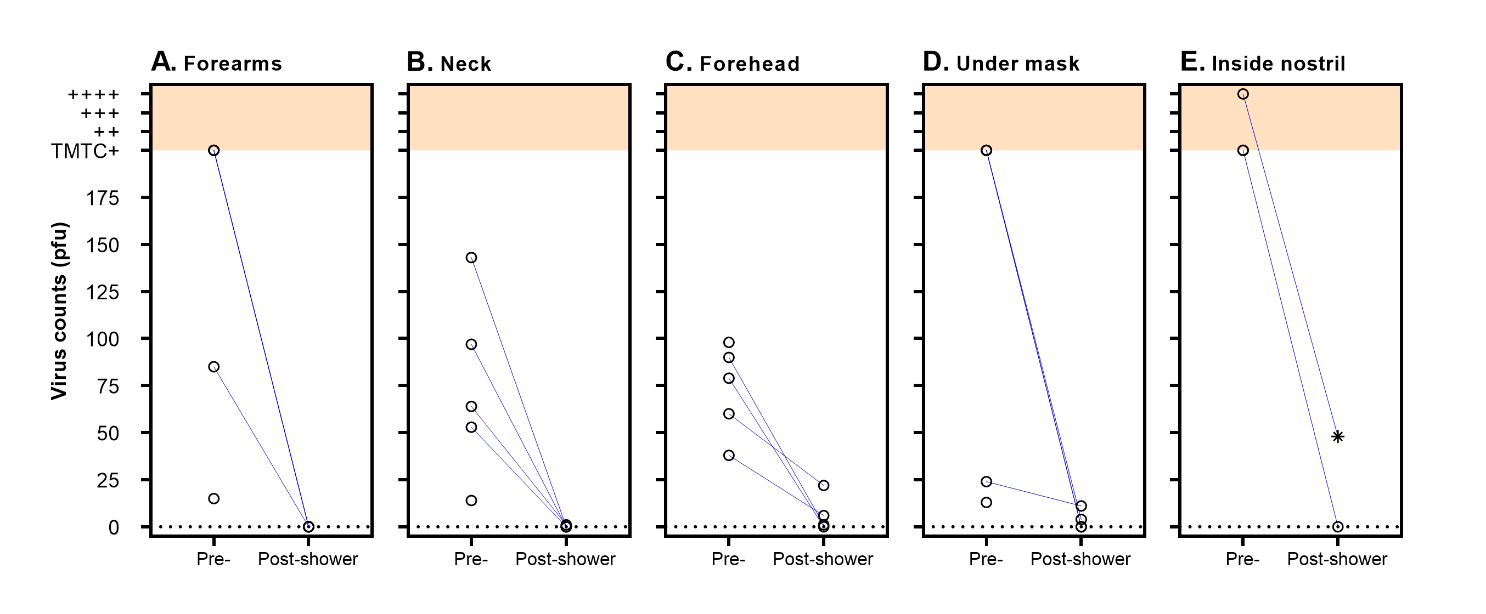

***Figure S6 – Virus counts pre- and post-shower for no PPE control (experiment 1).*** *Virus counts recovered from skin swabs (open circles, y-axis) are shown pre- (i.e. post virus aerosol exposure) and post-shower (x-axes). Virus counts were quantified as plaque forming units (PFU). Virus counts are shown relate to only those data from experiment 1 in the no PPE control condition. Note that there several data points where there is a missing post-shower sample (no connecting line). These cases correspond to the final condition tested on a day, where no post-shower samples were collected.*
